# African-specific improvement of a polygenic hazard score for age at diagnosis of prostate cancer

**DOI:** 10.1101/2020.04.20.20072926

**Authors:** Roshan Karunamuni, Minh-Phuong Huynh-Le, Chun Fan, Wesley Thompson, Rosalind Eeles, Zsofia Kote-Jarai, Kenneth Muir, UKGPCS collaborators, Artitaya Lophatananon, Catherine Tangen, Phyllis Goodman, Ian Thompson, William Blot, Wei Zheng, Adam Kibel, Bettina Drake, Olivier Cussenot, Geraldine Cancel-Tassin, Florence Menegaux, Therese Truong, Jong Park, Hui-Yi Lin, Jeannette Bensen, Elizabeth Fontham, James Mohler, Jack Taylor, Luc Multigner, Pascal Blanchet, Laurent Brureau, Marc Romana, Robin Leach, Esther John, Jay Fowke, William Bush, Melinda Aldrich, Dana Crawford, Shiv Srivastava, Jennifer Cullen, Gyorgy Petrovics, Marie-Elise Parent, Jennifer Hu, Maureen Sanderson, Ian Mills, Ole Andreassen, Anders Dale, Tyler M Seibert, The PRACTICAL Consortium

## Abstract

**Introduction:** Polygenic hazard score (PHS) models are associated with age at diagnosis of prostate cancer. Our model developed in Europeans (PHS46), showed reduced performance in men with African genetic ancestry. We used a cross-validated search to identify SNPs that might improve performance in this population.

**Material and Methods:** Anonymized genotypic data were obtained from the PRACTICAL consortium for 6,253 men with African genetic ancestry. Ten iterations of a ten-fold cross-validation search were conducted, to select SNPs that would be included in the final PHS46+African model. The coefficients of PHS46+African were estimated in a Cox proportional hazards framework using age at diagnosis as the dependent variable and PHS46, and selected SNPs as predictors. The performance of PHS46 and PHS46+African were compared using the same cross-validated approach.

**Results:** Three SNPs (rs76229939, rs74421890, and rs5013678) were selected for inclusion in PHS46+African. All three SNPs are located on chromosome 8q24. PHS46+African showed substantial improvements in all performance metrics measured, including a 75% increase in the relative hazard of those in the upper 20% compared to the bottom 20% (2.47 to 4.34) and a 20% reduction in the relative hazard of those in the bottom 20% compared to the middle 40% (0.65 to 0.53).

**Conclusions:** We identified three SNPs that substantially improved the association of PHS46 with age at diagnosis of prostate cancer in men with African genetic ancestry to levels comparable to Europeans and Asians. A strategy of building on established statistical models might benefit ancestral groups generally under-represented in genome-wide association studies.

## Introduction

Polygenic models can provide personalized estimates of the risk of developing prostate cancer. In the context of survival analysis, these models can provide insight into age at diagnosis of prostate cancer, and thus could be used to guide decisions on whether and when to offer screening^1^. Studies of polygenic models have often included only individuals of European genetic ancestry, owing to greater availability of data from that population^2,3^. As a consequence, these models have been tailored to identify and estimate coefficients of genetic common variants for that particular population, while potentially missing variants that may hold value in other populations^2^. There is concern that using these European-focused models could actually exacerbate health disparities^2–4^.

As an example, our group recently published on the performance of a polygenic hazard score (PHS) originally developed using a European dataset, in a multi-ethnic dataset consisting of individuals of European, African, and Asian genetic ancestry^5^. The model (called here PHS46), includes 46 single nucleotide polymorphisms (SNPs) in its calculation and was strongly associated with age at diagnosis in all three genetic populations (p<10^−16^). However, the hazard ratio for prostate cancer between individuals in the upper 20^th^ percentile to those in the lower 20^th^ percentile of PHS46 was approximately half as large for those with African genetic ancestry (2.6) as it was for those with European (5.6) or Asian (4.6) ancestry. A similar pattern was observed for clinically significant prostate cancer and for death from prostate cancer.

In the current study, we attempt to bridge the apparent gap in model performance of PHS46 for individuals with African genetic ancestry. To this end, we used a machine learning approach to systematically search for SNPs that add statistical value to a base model of PHS46 among African men (PHS46+African). By including PHS46 as a covariate in our SNP search, we sought to identify those SNPs that may hold particular value for individuals with African genetic ancestry.

## Material and Methods

### Study dataset

We obtained genotype and phenotype data from the Prostate Cancer Association Group to Investigate Cancer Associated Alterations in the Genome (PRACTICAL)^6^ consortium for this study. Genotyping was performed using the OncoArray platform^6^ and had undergone quality assurance steps, as described previously^7^. The genotypic ancestry of each individual was also determined previously^6,8^. In total, the African dataset consisted of data from 6,253 men with African genotypic ancestry. Missing SNP calls were replaced with the mean of the genotyped data for that SNP in the African dataset. Individuals without prostate cancer were censored at age at last follow-up in the Cox proportional hazards models. All contributing studies were approved by the relevant ethics committees; written informed consent was obtained from the study participants^9^. The present analyses used de-identified data from the PRACTICAL consortium. Please refer to Table S1 for a description of the PRACTICAL study groups that contributed data towards this analysis. PHS46 risk score for each individual in the African dataset was estimated as the sum of SNP allele counts (X) multiplied by their respective coefficients (β)^5^:

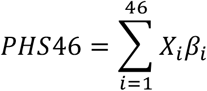

### SNP-scan

Training and testing sets were generated using 10 iterations of a 10-fold cross-validation structure resulting in 100 total permutations. For each permutation, a multivariable logistic regression model using case/control status as the dependent variable was estimated using each genotyped SNP in turn, adjusting for PHS46 and four principal components based on genetic ancestry, determined previously^8^. SNPs with p-values less than 1×10^−6^ were considered for further analysis. In order of increasing p-value, each SNP was tested in a multiple Cox proportional hazards model, after adjusting for PHS46, four ancestral principal components, and previously selected SNPs. The Cox model in the SNP-scan used age at diagnosis of prostate cancer as the dependent variable. If the p-value of the coefficient of the tested SNP was less than 1×10^−6^, it was considered for the final model in that permutation. SNPs that reached this p-value threshold in more than 50% of the permutations were selected to construct the PHS46+African model, consisting of PHS46 and the newly identified SNPs.

### Comparing performance between PHS46 and PHS46+African

For each permutation of the previously described cross-validation structure, an PHS46+African Cox proportional hazards model was estimated in the training set using PHS46 and the selected SNPs as independent predictors. The PHS46+African risk score for each individual is then estimated using the corresponding PHS46 score, selected SNP allele counts (Y) and their respective coefficients (α):

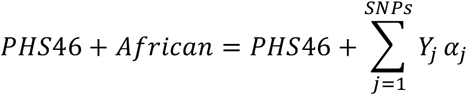

The performance of the PHS46+African and PHS46 models was then determined in the cross-validation testing set, and the resulting hazard ratios (HR) were obtained, as previously described^1^. For each model, the PHS risk scores within the cross-validation testing set are assigned to quantile groups identified using the corresponding training set control values. The hazard ratio between two quantile groups, such as those in the top 20% to those in the bottom 20%, is estimated as the exponential of the difference in mean PHS values for each group. In this calculation, the PHS values are linearly scaled by a sample-weight correction factor, to account for case-control sampling^1,5,10^. Three HR were calculated: HR80/20 (top 20% to bottom 20%), HR98/50 (top 2% to middle 40%) and HR20/50 (bottom 20% to middle 40%). The average HR across permutations for both PHS46+African and PHS46 are reported.

To allow for comparisons with previously published results, the performance metrics for PHS46 and PHS46+African were also estimated for age at diagnosis of clinically significant prostate cancer. When estimating performance for clinically significant prostate cancer, controls and non-clinically significant cancers were censored at age of last follow-up and age of diagnosis, respectively. The previously used criteria for clinically significant cancer were any of: Gleason score >=7, stage T3-T4, PSA concentration >= 10ng/mL, nodal metastasis, or distant metastasis^1^. Paired t-tests were used to test for statistical significant differences (α = 0.05) in HR between PHS46+African and PHS46.

Additionally, in each permutation, the performance of a Cox model consisting of PHS46 and SNPs that were considered in that permutation was also estimated. These results are provided within the Table S2 and provide performance estimates that are not prone to information leakage from training to testing set.

### Characterization of PHS+African

Coefficients of the PHS46+African model, consisting of PHS46 and the SNPs selected in the SNP-scan, were estimated using 1000 bootstrapped samples of the African dataset.

## Results

### Individual and OncoArray characteristics

In total, there were 3,013 men with (cases) and 3,240 men without (controls) prostate cancer in the African dataset. The mean [95% CI] ages of cases and controls were 62.4 [62.1, 62.7] and 61.8 [61.4, 62.1] years respectively. The OncoArray genotypic data, after the quality assurance process, included 444,323 SNPs.

### Single Nucleotide Polymorphism (SNP)-scan

Across the 100 permutations of the cross-validation iterations, a total of twelve SNPs were considered for final selection (Table S3). Three SNPs were selected in more than 50% of the permutations and included in the final PHS46+African model. By cross-referencing the chromosomal positions against dbSNP^11^, these variants were identified as rs76229939, rs74421890, and rs5013678. All 3 SNPs (Table 1) are located on chromosome 8q24, a region of the chromosome previously identified as containing common variants associated with prostate cancer^12,13^. An examination of the Pearson correlation coefficients (Table S4) showed little correlation, ranging from -0.05 to -0.07, among genotype data from the 3 SNPs in the African dataset.

**Table 1.** Characteristics of PHS46+African SNPs. RS-ID, chromosome and base-pair position (based on version 37), effect and reference alleles, bootstrap-estimated beta, and effect allele frequencies in Africans from 1000Genomes (referenced from dbSNP) of the three SNPs selected for addition to PHS46.

| RS number | Chromosome | Position | Effect | Ref | beta | Frequency (%) |
| --- | --- | --- | --- | --- | --- | --- |
| rs76229939 | 8 | 128085394 | G | A | 0.441 | 4.8 |
| rs74421890 | 8 | 128096183 | A | G | 0.415 | 4.1 |
| rs5013678 | 8 | 128103979 | G | A | -0.260 | 8.1 |

Reference threshold (Table S5) and mean (Table S6) values for PHS46+African in the African dataset are presented in the Supplemental Data.

### Performance of PHS46+African

Figure 1 demonstrates the difference in HRs between PHS46+African and PHS46 within the African dataset using age at diagnosis of any prostate cancer. Overall, we observed an improvement in all the metrics calculated: a 75% increase in HR98/50 from 2.10 to 3.67; a 79% increase in HR80/20 from 2.47 to 4.42; and a 23% decrease in HR20/50 from 0.65 to 0.51. We also observed improvements in all performance metrics when using age at diagnosis of clinically significant prostate cancer: 103% increase in HR98/50 from 1.91 to 3.88, 113% improvement in HR80/20 from 2.21 to 4.71, and 29% improvement in HR20/50 from 0.70 to 0.50. All observed changes in HR were statistically significant (p < 1×10^−16^).

**Figure 1.**
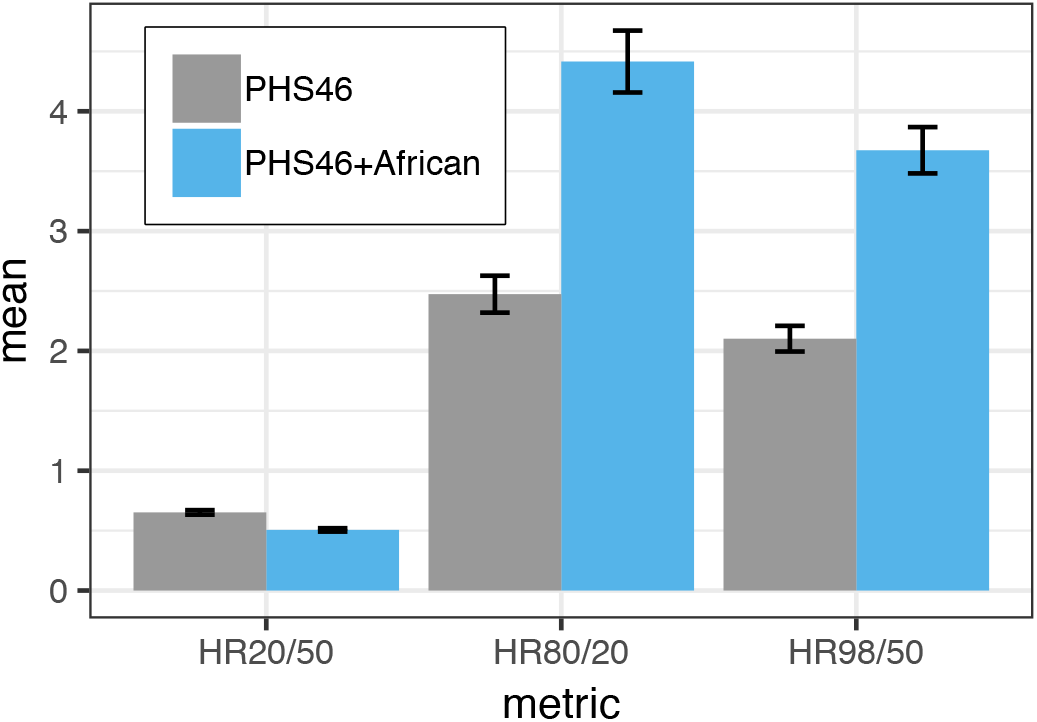
Comparison between PHS46 and PHS46+African. Mean hazard ratio metrics plotted for PHS46 and PHS46+African models in the African dataset. Improvements were observed in all performance metrics investigated. Error bars represent 95% confidence interval.

## Discussion

Using a cross-validated search of a dataset made up entirely of men with African genetic ancestry, we were able to identify three SNPs that substantially improved the performance of PHS46 in this population to levels that are comparable to those observed in Europeans and Asians. The three SNPs, rs76229939, rs74421890, and rs5013678, are all located on chromosome 8q24 – a region of the genome where variants have been associated with prostate cancer in both the general population and specifically in men with African genetic ancestry^13,14^. Despite the relative proximity of the three SNPs on chromosome 8, their genetic data was not strongly correlated in our dataset, suggesting that each SNP provides non-redundant information for an individual’s genetic score.

Each of the three SNPs have been previously identified in the literature to be associated with prostate cancer: rs76229939 is an intron variant of the prostate-cancer-associated transcript 2 (PCAT2) gene, while rs74421890 and rs5013678 are both non-coding transcript variants of the prostate-cancer-associated non-coding RNA 1 (PRNCR1) gene. The minor allele frequencies of rs76229939 and rs74421890 in Europeans, as reported by dbSNP^11^, are approximately zero to three decimal places, which may explain why they were not selected in the original formulation of PHS46.

This study is not meant to be an exhaustive search for all possible SNPs that are associated with the age of diagnosis of prostate cancer in individuals with African genetic ancestry. Our study is also limited by the small number of available observations relative to those often found in many genome-wide association studies, which can have tens or hundreds of thousands of individuals. However, we were able to extract information that is likely robust by employing a cross-validated search for those SNPs that specifically add value to the performance of PHS46, and not simply independently associated with prostate cancer. We also note that no SNP score, including PHS46 and PHS46+African, has been shown to discriminate men at risk of aggressive prostate cancer from those at risk of indolent prostate cancer. Finally, the performance metrics reported in this study may be biased by the leakage of information across cross-validated folds of the data when identifying those SNPs to include in the final African-PHS model. This bias is expected to be similar for all SNPs and should not have influenced selection of the three SNPs included in the final model over those not selected.

In conclusion, we identified three SNPs (rs76229939, rs74421890, and rs5013678) that substantially improved the performance of PHS46 in a dataset of men with African genetic ancestry. We believe that this strategy of building on established models developed on large dataset could be applied to other groups that are generally under-represented in genome-wide association studies. This strategy may further help to bridge performance gaps in personalized genetic risk scores across populations.

## Data Availability

The data used in this work were obtained from the Prostate Cancer Association Group to Investigate Cancer Associated Alterations in the Genome (PRACTICAL) consortium,
Readers who are interested in accessing the data must first submit a proposal to the Data Access Committee. If the reader is not a member of the consortium, their concept form must be sponsored by a principal investigator (PI) of one of the PRACTICAL consortium member studies. If approved by the Data Access Committee, PIs within the consortium, each of whom retains ownership of their data submitted to the consortium, can then choose to participate in the specific proposal. In addition, portions of the data are available for request from dbGaP (database of Genotypes and Phenotypes) which is maintained by the National Center for Biotechnology Information (NCBI): https://www.ncbi.nlm.nih.gov/gap/?term=Icogs+prostatehttps://www.ncbi.nlm.nih.gov/gap/?term=Icogs+prostate.
Anyone can apply to join the consortium. The eligibility requirements are listed here: http://practical.icr.ac.uk/blog/?page_id=9. Joining the consortium would not guarantee access, as a proposal for access would still be submitted to the Data Access Committee, but there would be no need for a separate member sponsor. Readers may find information about application by using the contact information below:
Rosalind Eeles
Principal Investigator for PRACTICAL
Professor of Oncogenetics
Institute of Cancer Research (ICR)
Sutton, UK
URL: http://practical.icr.ac.uk
Tel: ++44 (0)20 8722 4094

## Supplemental Data Description

The Supplemental Data contains (1) six tables

## Appendix A1. Data Availability Statement

The data used in this work were obtained from the Prostate Cancer Association Group to Investigate Cancer Associated Alterations in the Genome (PRACTICAL) consortium, Readers who are interested in accessing the data must first submit a proposal to the Data Access Committee. If the reader is not a member of the consortium, their concept form must be sponsored by a principal investigator (PI) of one of the PRACTICAL consortium member studies. If approved by the Data Access Committee, PIs within the consortium, each of whom retains ownership of their data submitted to the consortium, can then choose to participate in the specific proposal. In addition, portions of the data are available for request from dbGaP (database of Genotypes and Phenotypes) which is maintained by the National Center for Biotechnology Information (NCBI): https://www.ncbi.nlm.nih.gov/gap/?term=Icogs+prostatehttps://www.ncbi.nlm.nih.gov/gap/?term=Icogs+prostate. Anyone can apply to join the consortium. The eligibility requirements are listed here: http://practical.icr.ac.uk/blog/?page_id=9. Joining the consortium would not guarantee access, as a proposal for access would still be submitted to the Data Access Committee, but there would be no need for a separate member sponsor. Readers may find information about application by using the contact information below:

Rosalind Eeles

Principal Investigator for PRACTICAL

Professor of Oncogenetics

Institute of Cancer Research (ICR)

Sutton, UK

URL: http://practical.icr.ac.uk

Tel: ++44 (0)20 8722 4094

## Appendix A2. Funding sources for the PRACTICAL consortium

### CRUK and PRACTICAL consortium

This work was supported by the Canadian Institutes of Health Research, European Commission’s Seventh Framework Programme grant agreement n° 223175 (HEALTH-F2-2009-223175), Cancer Research UK Grants C5047/A7357, C1287/A10118, C1287/A16563, C5047/A3354, C5047/A10692, C16913/A6135, and The National

Institute of Health (NIH) Cancer Post-Cancer GWAS initiative grant: No. 1 U19 CA 148537-01 (the GAME-ON initiative).

We would also like to thank the following for funding support: The Institute of Cancer Research and The Everyman Campaign, The Prostate Cancer Research Foundation, Prostate Research Campaign UK (now Prostate Action), The Orchid Cancer Appeal, The National Cancer Research Network UK, The National Cancer Research Institute (NCRI) UK. We are grateful for support of NIHR funding to the NIHR Biomedical Research Centre at The Institute of Cancer Research and The Royal Marsden NHS Foundation Trust.

The Prostate Cancer Program of Cancer Council Victoria also acknowledge grant support from The National Health and Medical Research Council, Australia (126402, 209057, 251533,, 396414, 450104, 504700, 504702, 504715, 623204, 940394, 614296,), VicHealth, Cancer Council Victoria, The Prostate Cancer Foundation of Australia, The Whitten Foundation, PricewaterhouseCoopers, and Tattersall’s. EAO, DMK, and EMK acknowledge the Intramural Program of the National Human Genome Research Institute for their support.

Genotyping of the OncoArray was funded by the US National Institutes of Health (NIH) [U19 CA 148537 for ELucidating Loci Involved in Prostate cancer SuscEptibility (ELLIPSE) project and X01HG007492 to the Center for Inherited Disease Research (CIDR) under contract number HHSN268201200008I].

This study would not have been possible without the contributions of the following: Coordination team, bioinformatician and genotyping centers: Genotyping at CCGE, Cambridge: Caroline Baines and Don Conroy

Funding for the iCOGS infrastructure came from: the European Community’s Seventh Framework Programme under grant agreement n° 223175 (HEALTH-F2-2009-223175) (COGS), Cancer Research UK (C1287/A10118, C1287/A 10710, C12292/A11174, C1281/A12014, C5047/A8384, C5047/A15007, C5047/A10692, C8197/A16565), the National Institutes of Health (CA128978) and Post-Cancer GWAS initiative (1U19 CA148537, 1U19 CA148065 and 1U19 CA148112 - the GAME-ON initiative), the Department of Defence (W81XWH-10-1-0341), the Canadian Institutes of Health Research (CIHR) for the CIHR Team in Familial Risks of Breast Cancer, Komen Foundation for the Cure, the Breast Cancer Research Foundation, and the Ovarian Cancer Research Fund.

This study would not have been possible without the contributions of the following: Per Hall (COGS); Douglas F. Easton, Paul Pharoah, Kyriaki Michailidou, Manjeet K. Bolla, Qin Wang (BCAC), Andrew Berchuck (OCAC), Rosalind A. Eeles, Douglas F. Easton, Ali Amin Al Olama, Zsofia Kote-Jarai, Sara Benlloch (PRACTICAL), Georgia Chenevix-Trench, Antonis Antoniou, Lesley McGuffog, Fergus Couch and Ken Offit (CIMBA), Joe Dennis, Alison M. Dunning, Andrew Lee, and Ed Dicks, Craig Luccarini and the staff of the Centre for Genetic Epidemiology Laboratory, Javier Benitez, Anna Gonzalez-Neira and the staff of the CNIO genotyping unit, Jacques Simard and Daniel C. Tessier, Francois Bacot, Daniel Vincent, Sylvie LaBoissière and Frederic Robidoux and the staff of the McGill University and Génome Québec Innovation Centre, Stig E. Bojesen, Sune F. Nielsen, Borge G. Nordestgaard, and the staff of the Copenhagen DNA laboratory, and Julie M. Cunningham, Sharon A. Windebank, Christopher A. Hilker, Jeffrey Meyer and the staff of Mayo Clinic Genotyping Core Facility

Additional funding and acknowledgments from studies in PRACTICAL:

Information of the PRACTICAL consortium can be found at http://practical.icr.ac.uk/

### BioVU

The dataset(s) used for the analyses described were obtained from Vanderbilt University Medical Center’s BioVU, which is supported by institutional funding and by the National Center for Research Resources, Grant UL1 RR024975-01 (which is now at the National Center for Advancing Translational Sciences, Grant 2 UL1 TR000445-06).

### CPDR

Uniformed Services University for the Health Sciences HU0001-10-2-0002 (PI: David G. McLeod, MD)

### EPICAP

The EPICAP study was supported by grants from Ligue Nationale Contre le Cancer; Institut National du Cancer (INCa); Fondation ARC; Fondation de France; Agence Nationale de sécurité sanitaire de l’alimentation, de l’environnement et du travail (ANSES); Ligue départementale du Val de Marne. The EPICAP study group would like to thank all urologists, Antoinette Anger and Hasina Randrianasolo (study monitors), Anne-Laure Astolfi, Coline Bernard, Oriane Noyer, Marie-Hélène De Campo, Sandrine Margaroline, Louise N’Diaye, Sabine Perrier-Bonnet (Clinical Research nurses)

### KARUPROSTATE

The Karuprostate study was supported by the the Frech National Health Directorate, the Association pour la Recherche sur le Cancer, la Ligue Nationale contre le Cancer, the French Agency for Environmental and Occupational Health Safety (ANSES) and by the Association pour la Recherche sur les Tumeurs de la Prostate. We would like to thank Séverine Ferdinand for valuable contributions to the study.

### MOFFITT

The Moffitt group was supported by the US National Cancer Institute (R01CA128813, PI: J.Y. Park).

### NMHS

Funding for the Nashville Men’s Health Study (NMHS) was provided by the National Institutes of Health Grant numbers: RO1CA121060

### PCaP

The North Carolina - Louisiana Prostate Cancer Project (PCaP) and the Health Care Access and Prostate Cancer Treatment in North Carolina (HCaP-NC) study are carried out as collaborative studies supported by the Department of Defense contract DAMD 17-03-2-0052 and the American Cancer Society award RSGT-08-008-01-CPHPS, respectively.

The authors thank the staff, advisory committees and research subjects participating in the PCaP and HCaP-NC studies for their important contributions.

### PROtEuS

PROtEuS was supported financially through grants from the Canadian Cancer Society [13149, 19500, 19864, 19865] and the Cancer Research Society, in partnership with the Ministère de l’enseignement supérieur, de la recherche, de la science et de la technologie du Québec, and the Fonds de la recherche du Québec - Santé.PROtEuS would like to thank its collaborators and research personnel, and the urologists involved in subjects recruitment. We also wish to acknowledge the special contribution made by Ann Hsing and Anand Chokkalingam to the conception of the genetic component of PROtEuS.

### SABOR

The SABOR research is supported by NIH/NCI Early Detection Research Network, grant U01 CA0866402-18. Also supported by the Cancer Center Support Grant to the Mays Cancer Center from the National Cancer Institute (US) P30 CA054174

### SCCS

SCCS is funded by NIH grant R01 CA092447, and SCCS sample preparation was conducted at the Epidemiology Biospecimen Core Lab that is supported in part by the Vanderbilt-Ingram Cancer Center (P30 CA68485). Data on SCCS cancer cases used in this publication were provided by the Alabama Statewide Cancer Registry; Kentucky Cancer Registry, Lexington, KY; Tennessee Department of Health, Office of Cancer Surveillance; Florida Cancer Data System; North Carolina Central Cancer Registry, North Carolina Division of Public Health; Georgia Comprehensive Cancer Registry; Louisiana Tumor Registry; Mississippi Cancer Registry; South Carolina Central Cancer Registry; Virginia Department of Health, Virginia Cancer Registry; Arkansas Department of Health, Cancer Registry, 4815 W. Markham, Little Rock, AR 72205. The Arkansas Central Cancer Registry is fully funded by a grant from National Program of Cancer Registries, Centers for Disease Control and Prevention (CDC). Data on SCCS cancer cases from Mississippi were collected by the Mississippi Cancer Registry which participates in the National Program of Cancer Registries (NPCR) of the Centers for Disease Control and Prevention (CDC). The contents of this publication are solely the responsibility of the authors and do not necessarily represent the official views of the CDC or the Mississippi Cancer Registry.

### SCPCS

SCPCS is funded by CDC grant S1135-19/19, and SCPCS sample preparation was conducted at the Epidemiology Biospecimen Core Lab that is supported in part by the Vanderbilt-Ingram Cancer Center (P30 CA68485).

### SFPCS

SFPCS was funded by California Cancer Research Fund grant 99-00527V-10182

### SWOG-PCPT / SWOG-SELECT

PCPT and SELECT are funded by Public Health Service grants U10CA37429 and 5UM1CA182883 from the National Cancer Institute. The authors thank the site investigators and staff and, most importantly, the participants from PCPT who donated their time to this trial.

### UKGPCS

UKGPCS would also like to thank the following for funding support: The Institute of Cancer Research and The Everyman Campaign, The Prostate Cancer Research Foundation, Prostate Research Campaign UK (now Prostate Action), The Orchid Cancer Appeal, The National Cancer Research Network UK, The National Cancer Research Institute (NCRI) UK. We are grateful for support of NIHR funding to the NIHR Biomedical Research Centre at The Institute of Cancer Research and The Royal Marsden NHS Foundation Trust. UKGPCS should also like to acknowledge the NCRN nurses, data managers and Consultants for their work in the UKGPCS study.UKGPCS would like to thank all urologists and other persons involved in the planning, coordination, and data collection of the study. KM and AL were in part supported from the NIHR Manchester Biomedical Research Centre

### WUGS

WUGS would like to thank the following for funding support: The Anthony DeNovi Fund, the Donald C. McGraw Foundation, and the St. Louis Men’s Group Against Cancer.

## Appendix A3 Members of the PRACTICAL Consortium

Christopher A. Haiman^1^, Fredrick R. Schumacher^2,3^, Sara Benlloch^4,5^, Ali Amin Al Olama^6,7^, Sonja I. Berndt^8^, David V. Conti^1^, Fredrik Wiklund^9^, Stephen Chanock^8^, Susan M. Gapstur^10^, Victoria L. Stevens^10^, Jyotsna Batra^11,12^, Judith Clements^11,12^, APCB BioResource^13,14^, Henrik Grönberg^15^, Nora Pashayan^16,17^, Johanna Schleutker^18,19^, Demetrius Albanes^8^, Stephanie Weinstein^8^, Alicja Wolk^20,21^, Catharine West^22^, Lorelei Mucci^23^, Stella Koutros^8^, Karina Dalsgaard Sørensen^24,25^, Eli Marie Grindedal^26^, David E. Neal^27,28,29^, Freddie C. Hamdy^30,31^, Jenny L. Donovan^32^, Ruth C. Travis^33^, Robert J. Hamilton^34,35^, Sue Ann Ingles^36^, Barry S. Rosenstein^37,38^, Yong-Jie Lu^39^, Graham G. Giles^40,41,42^, Ana Vega^43,44,45^, Manolis Kogevinas^46,47,48,49^, Kathryn L. Penney^50^, Janet L. Stanford^51,52^, Cezary Cybulski^53^, Børge G. Nordestgaard^54,55^, Hermann Brenner^56,57,58^, Christiane Maier^59^, Jeri Kim^60^, Manuel R. Teixeira^61,62^, Susan L. Neuhausen^63^, Kim De Ruyck^64^, Azad Razack^65^, Lisa F. Newcomb^51,66^, Davor Lessel^67^, Radka Kaneva^68^, Nawaid Usmani^69,70^, Frank Claessens^71^, Paul A. Townsend^72^, Manuela Gago-Dominguez^73,74^, Monique J. Roobol^75^, Kay-Tee Khaw^76^, Lisa Cannon-Albright^77,78^, Hardev Pandha^79^, Stephen N. Thibodeau^80^, Peter Kraft^81^, Elio Riboli^82^

^1^Center for Genetic Epidemiology, Department of Preventive Medicine, Keck School of Medicine, University of Southern California/Norris Comprehensive Cancer Center, Los Angeles, CA 90015, USA

^2^Department of Population and Quantitative Health Sciences, Case Western Reserve University, Cleveland, OH 44106-7219, USA

^3^Seidman Cancer Center, University Hospitals, Cleveland, OH 44106, USA.

^4^Centre for Cancer Genetic Epidemiology, Department of Public Health and Primary Care, University of Cambridge, Strangeways Research Laboratory, Cambridge CB1 8RN, UK

^5^The Institute of Cancer Research, London, SM2 5NG, UK

^6^Centre for Cancer Genetic Epidemiology, Department of Public Health and Primary Care, University of Cambridge, Strangeways Research Laboratory, Cambridge, UK

^7^University of Cambridge, Department of Clinical Neurosciences, Stroke Research Group, R3, Box 83, Cambridge Biomedical Campus, Cambridge CB2 0QQ, UK

^8^Division of Cancer Epidemiology and Genetics, National Cancer Institute, NIH, Bethesda, Maryland, 20892, USA

^9^Department of Medical Epidemiology and Biostatistics, Karolinska Institute, SE-171 77 Stockholm, Sweden

^10^Behavioral and Epidemiology Research Group, Research Program, American Cancer Society, 250 Williams Street, Atlanta, GA 30303, USA

^11^Australian Prostate Cancer Research Centre-Qld, Institute of Health and Biomedical Innovation and School of Biomedical Sciences, Queensland University of Technology, Brisbane QLD 4059, Australia

^12^Translational Research Institute, Brisbane, Queensland 4102, Australia

^13^Australian Prostate Cancer Research Centre-Qld, Queensland University of Technology, Brisbane; Prostate Cancer Research Program, Monash University, Melbourne; Dame Roma Mitchell Cancer Centre, University of Adelaide, Adelaide; Chris O’Brien Lifehouse and

^14^Translational Research Institute, Brisbane, Queensland, Australia

^15^Department of Medical Epidemiology and Biostatistics, Karolinska Institute, Stockholm, Sweden

^16^Department of Applied Health Research, University College London, London, WC1E 7HB, UK

^17^Centre for Cancer Genetic Epidemiology, Department of Oncology, University of Cambridge, Strangeways Laboratory, Worts Causeway, Cambridge, CB1 8RN, UK

^18^Institute of Biomedicine, Kiinamyllynkatu 10, FI-20014 University of Turku, Finland

^19^Department of Medical Genetics, Genomics, Laboratory Division, Turku University Hospital, PO Box 52, 20521 Turku, Finland

^20^Division of Nutritional Epidemiology, Institute of Environmental Medicine, Karolinska Institutet, SE-171 77 Stockholm, Sweden

^21^Department of Surgical Sciences, Uppsala University, 75185 Uppsala, Sweden

^22^Division of Cancer Sciences, University of Manchester, Manchester Academic Health Science Centre, Radiotherapy Related Research, The Christie Hospital NHS Foundation Trust, Manchester, M13 9PL UK

^23^Department of Epidemiology,Harvard T. H. Chan School of Public Health, Boston, MA 02115, USA

^24^Department of Molecular Medicine, Aarhus University Hospital, Palle Juul-Jensen Boulevard 99, 8200 Aarhus N, Denmark

^25^Department of Clinical Medicine, Aarhus University, DK-8200 Aarhus N

^26^Department of Medical Genetics, Oslo University Hospital, 0424 Oslo, Norway

^27^Nuffield Department of Surgical Sciences, University of Oxford, Room 6603, Level 6, John Radcliffe Hospital, Headley Way, Headington, Oxford, OX3 9DU, UK

^28^University of Cambridge, Department of Oncology, Box 279, Addenbrooke’s Hospital, Hills Road, Cambridge CB2 0QQ, UK

^29^Cancer Research UK, Cambridge Research Institute, Li Ka Shing Centre, Cambridge UK

^30^Nuffield Department of Surgical Sciences, University of Oxford, Oxford, OX1 2JD, UK

^31^Faculty of Medical Science, University of Oxford, John Radcliffe Hospital, Oxford, UK

^32^Population Health Sciences, Bristol Medical School, University of Bristol, BS8 2PS, UK

^33^Cancer Epidemiology Unit, Nuffield Department of Population Health, University of Oxford, Oxford, OX3 7LF, UK

^34^Dept. of Surgical Oncology, Princess Margaret Cancer Centre, Toronto ON M5G 2M9, Canada

^35^Dept. of Surgery (Urology), University of Toronto, Canada

^36^Department of Preventive Medicine, Keck School of Medicine, University of Southern California/Norris Comprehensive Cancer Center, Los Angeles, CA 90015, USA

^37^Department of Radiation Oncology and Department of Genetics and Genomic Sciences, Box 1236, Icahn School of Medicine at Mount Sinai, One Gustave L. Levy Place, New York, NY 10029, USA

^38^Department of Genetics and Genomic Sciences, Icahn School of Medicine at Mount Sinai, New York, NY 10029-5674, USA.

^39^Centre for Molecular Oncology, Barts Cancer Institute, Queen Mary University of London, John Vane Science Centre, Charterhouse Square, London, EC1M 6BQ, UK

^40^Cancer Epidemiology Division, Cancer Council Victoria, 615 St Kilda Road, Melbourne, VIC 3004, Australia

^41^Centre for Epidemiology and Biostatistics, Melbourne School of Population and Global Health, The University of Melbourne, Grattan Street, Parkville, VIC 3010, Australia

^42^Precision Medicine, School of Clinical Sciences at Monash Health, Monash University, Clayton, Victoria 3168, Australia

^43^Fundación Pública Galega Medicina Xenómica, Santiago De Compostela, 15706, Spain.

^44^Instituto de Investigación Sanitaria de Santiago de Compostela, Santiago De Compostela, 15706, Spain.

^45^Centro de Investigación en Red de Enfermedades Raras (CIBERER), Spain

^46^ISGlobal, Barcelona, Spain

^47^IMIM (Hospital del Mar Medical Research Institute), Barcelona, Spain

^48^Universitat Pompeu Fabra (UPF), Barcelona, Spain

^49^CIBER Epidemiología y Salud Pública (CIBERESP), Madrid, Spain

^50^Channing Division of Network Medicine, Department of Medicine, Brigham and Women’s Hospital/Harvard Medical School, Boston, MA 02184, USA

^51^Division of Public Health Sciences, Fred Hutchinson Cancer Research Center, Seattle, Washington, 98109-1024, USA

^52^Department of Epidemiology, School of Public Health, University of Washington, Seattle, Washington 98195, USA

^53^International Hereditary Cancer Center, Department of Genetics and Pathology, Pomeranian Medical University, Szczecin, Poland

^54^Faculty of Health and Medical Sciences, University of Copenhagen, 2200 Copenhagen, Denmark

^55^Department of Clinical Biochemistry, Herlev and Gentofte Hospital, Copenhagen University Hospital, Herlev, 2200 Copenhagen, Denmark

^56^Division of Clinical Epidemiology and Aging Research, German Cancer Research Center (DKFZ), D-69120, Heidelberg, Germany

^57^German Cancer Consortium (DKTK), German Cancer Research Center (DKFZ), D-69120 Heidelberg, Germany

^58^Division of Preventive Oncology, German Cancer Research Center (DKFZ) and National Center for Tumor Diseases (NCT), Im Neuenheimer Feld 46069120 Heidelberg, Germany

^59^Humangenetik Tuebingen, Paul-Ehrlich-Str 23, D-72076 Tuebingen, Germany

^60^The University of Texas M. D. Anderson Cancer Center, Department of Genitourinary Medical Oncology, 1515 Holcombe Blvd., Houston, TX 77030, USA

^61^Department of Genetics, Portuguese Oncology Institute of Porto (IPO-Porto), Porto, Portugal

^62^Biomedical Sciences Institute (ICBAS), University of Porto, Porto, Portugal

^63^Department of Population Sciences, Beckman Research Institute of the City of Hope, 1500 East Duarte Road, Duarte, CA 91010, 626-256-HOPE (4673)

^64^Ghent University, Faculty of Medicine and Health Sciences, Basic Medical Sciences, Proeftuinstraat 86, B-9000 Gent

^65^Department of Surgery, Faculty of Medicine, University of Malaya, 50603 Kuala Lumpur, Malaysia

^66^Department of Urology, University of Washington, 1959 NE Pacific Street, Box 356510, Seattle, WA 98195, USA

^67^Institute of Human Genetics, University Medical Center Hamburg-Eppendorf, D-20246 Hamburg, Germany

^68^Molecular Medicine Center, Department of Medical Chemistry and Biochemistry, Medical University of Sofia, Sofia, 2 Zdrave Str., 1431 Sofia, Bulgaria

^69^Department of Oncology, Cross Cancer Institute, University of Alberta, 11560 University Avenue, Edmonton, Alberta, Canada T6G 1Z2

^70^Division of Radiation Oncology, Cross Cancer Institute, 11560 University Avenue, Edmonton, Alberta, Canada T6G 1Z2

^71^Molecular Endocrinology Laboratory, Department of Cellular and Molecular Medicine, KU Leuven, BE-3000, Belgium

^72^Division of Cancer Sciences, Manchester Cancer Research Centre, Faculty of Biology, Medicine and Health, Manchester Academic Health Science Centre, NIHR Manchester Biomedical Research Centre, Health Innovation Manchester, Univeristy of Manchester, M13 9WL

^73^Genomic Medicine Group, Galician Foundation of Genomic Medicine, Instituto de Investigacion Sanitaria de Santiago de Compostela (IDIS), Complejo Hospitalario Universitario de Santiago, Servicio Galego de Saúde, SERGAS, 15706, Santiago de Compostela, Spai

^74^University of California San Diego, Moores Cancer Center, La Jolla, CA 92037, USA

^75^Department of Urology, Erasmus University Medical Center, 3015 CE Rotterdam, The Netherlands

^76^Clinical Gerontology Unit, University of Cambridge, Cambridge, CB2 2QQ, UK

^77^Division of Epidemiology, Department of Internal Medicine, University of Utah School of Medicine, Salt Lake City, Utah, USA

^78^George E. Wahlen Department of Veterans Affairs Medical Center, Salt Lake City, Utah 84148, USA

^79^The University of Surrey, Guildford, Surrey, GU2 7XH

^80^Department of Laboratory Medicine and Pathology, Mayo Clinic, Rochester, MN 55905, USA

^81^Program in Genetic Epidemiology and Statistical Genetics, Department of Epidemiology, Harvard School of Public Health, Boston, MA, USA

^82^Department of Epidemiology and Biostatistics, School of Public Health, Imperial College London, SW7 2AZ, UK

**Table S1.** Contributing studies. Descriptions of PRACTICAL study groups that contributed data towards this analysis. The number of cases and controls provided by each study group is also listed.

| Study Group<br>Acronym | Study Group Name | Cases | Controls |
| --- | --- | --- | --- |
| BioVU | Vanderbilt University | 204 | 0 |
| CPDR | Uniformed Services University-Center for<br>Prostate Disease Research | 135 | 41 |
| CeRePP | French Prostate Case-Control Study | 101 | 84 |
| EPICAP | EPIdemiology of Prostate CAncer | 20 | 9 |
| KARUPROSTATE | French West Indies Prostate cancer Study | 363 | 386 |
| MIAMI-WFPCS | The University of Miami – Sylvester<br>Comprehensive Cancer Center | 59 | 49 |
| MOFFITT | The Moffitt Group | 101 | 91 |
| NMHS | Nashville Men’s Health Study | 176 | 188 |
| PCaP | North Carolina – Louisiana Prostate Cancer<br>Project Consortium | 967 | 0 |
| PROtEuS | Prostate Cancer and Environment Study | 70 | 53 |
| SABOR | San Antonio Center of Biomarkers of Risk for<br>Prostate Cancer | 105 | 106 |
| SCCS | Southern Community Cohort Study | 291 | 1498 |
| SCPCS | South Carolina Prostate Cancer Study | 57 | 32 |
| SFPCS | San Francisco Bay Area Prostate Cancer Study | 81 | 36 |
| SWOG-PCPT | Prostate Cancer Prevention Trial | 43 | 121 |
| SWOG-SELECT | Selenium and Vitamin E Cancer Prevention Trial | 30 | 167 |
| UKGPCS | U.K. Genetic Prostate Cancer Study and The<br>Prostate Cancer Research Foundation Study | 365 | 0 |
| WUGS | Washington University Genetics Study | 72 | 152 |

**Table S2.** Permutation performance. Mean and 95% confidence intervals for performance metrics estimated for each permutation of the cross-validation procedure.

| **metric** | **Any prostate cancer** | **Aggressive prostate cancer** |
| --- | --- | --- |
| HR20/50 | 0.53 [0.51-0.54] | 0.51 [0.50-0.53] |
| HR80/20 | 4.07 [3.81-4.32] | 4.42 [4.09-4.77] |
| HR98/50 | 3.49 [3.29-3.69] | 3.77 [3.50-4.04] |

**Table S3.** Results of SNP-scan. Description of 12 SNPs identified in 100 permutations of cross-validation SNP-scan. The position of each SNP is based on version 37. The count is the number of times the SNP appeared in permutations.

| RS number | Chromosome | Effect | Reference | Position | count |
| --- | --- | --- | --- | --- | --- |
| rs76229939 | 8 | G | A | 128085394 | 87 |
| rs74421890 | 8 | A | G | 128096183 | 87 |
| rs5013678 | 8 | G | A | 128103979 | 54 |
| rs144732329 | 2 | C | A | 20130787 | 18 |
| rs76595456 | 8 | A | G | 128087829 | 13 |
| rs339353 | 6 | A | C | 117202475 | 8 |
| rs1456315 | 8 | A | G | 128103937 | 6 |
| rs184167671 | 17 | G | A | 54159621 | 5 |
| rs339359 | 6 | A | G | 117160693 | 3 |
| rs610424 | 6 | G | A | 117212258 | 1 |
| rs339302 | 6 | T | A | 117224641 | 1 |
| rs6983561 | 8 | C | A | 128106880 | 1 |

**Table S4.** SNP correlation matrix. Pearson correlation coefficient matrix of three SNPs selected for addition to the PHS46+African model.

| RS number | rs76229939 | rs74421890 | rs5013678 |
| --- | --- | --- | --- |
| rs76229939 | 1 |  |  |
| rs74421890 | -0.0520 | 1 |  |
| rs5013678 | -0.0646 | -0.0759 | 1 |

**Table S5.** Reference Threshold PHS46+African scores. Reference threshold values for PHS46+African scores in African dataset.

| Threshold | Value |
| --- | --- |
| 20<sup>th</sup> percentile | -0.58 |
| 30<sup>th</sup> percentile | -0.47 |
| 70<sup>th</sup> percentile | -0.13 |
| 80<sup>th</sup> percentile | -0.02 |
| 98<sup>th</sup> percentile | 0.39 |

**Table S6.**
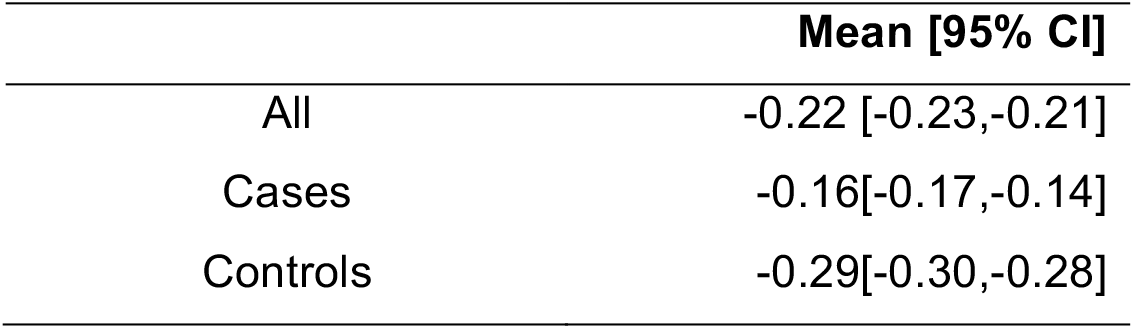
Mean PHS46+African scores. Mean PHS46+African risk scores are tabulated for all individuals, cases, and controls in the African dataset.

|  | Mean [95% CI] |
| --- | --- |
| All | -0.22 [-0.23,-0.21] |
| Cases | -0.16[-0.17,-0.14] |
| Controls | -0.29[-0.30,-0.28] |

## Notes

### Competing Interest Statement

The authors have declared no competing interest.

### Funding Statement

No external funding was received in support of the work presented.

